# The impact of a SmartPhone applicatiOn for skin cancer risk assessmenT on the healthcare system (SPOT-study): A randomized controlled trial

**DOI:** 10.1101/2025.11.18.25340297

**Authors:** Anna M Smak Gregoor, Tobias E Sangers, Carin A Uyl-de Groot, Eveline A M Heijnsdijk, Tamar EC Nijsten, Marlies Wakkee

## Abstract

**Background:** Artificial intelligence (AI)-based mobile health (mHealth) smartphone apps for skin cancer detection are increasingly available to the general population, but their impact on care is unclear.

**Methods:** The SPOT study is an investigator-initiated and -designed, unblinded, randomized controlled trial. Participants from a Dutch non-profit health insurance living in and around region Rotterdam the Netherlands, were recruited between August and December 2021. Participants were randomly assigned (3:2) to either free access to an AI-based mHealth app for skin cancer risk detection or care-as-usual. The primary endpoint was the difference in healthcare consumption for (pre)malignant and benign skin lesions at 12-months follow-up in the intention-to-treat population. Secondary endpoints included differences in the proportion of surgical interventions, overall use of dermatological care, and costs.

**Findings:** Among the 19,009 participants, the incidence of claims for (pre)malignant skin lesions was 2·8-fold higher than among non-responders. Within the group of study participants, the skin cancer incidence was higher among the intervention group compared to the control group at 12 months follow-up (2·7% (n=305) vs. 2·3% (n=171); risk difference (RD) 0·4% (95% confidence interval (CI) -0·07 to 0·85), p = 0·10), though this difference was not statistically significant. Furthermore, participants in the intervention group had significantly more claims for benign skin lesions (3·9% (n=443) vs. 2·6% (n=198), RD 1·3 (95% CI 0·7 to 1·7), p < 0·001), underwent more surgical interventions, and had higher mean costs per participant (€63 (95% CI 58 to -67), vs. €47 (41 to -52); p<0·001) compared to controls.

**Interpretation:** In the first 12 months of this study, access to an AI-based mHealth app for skin cancer risk detection showed a modest trend toward a higher rate of skin cancer detection compared to care-as-usual. However, it also resulted in significantly more dermatological care for benign skin lesions.

**Funding:** DSW and SkinVision®

**Research in context:** 

**Evidence before this study:** Prior to the start of this study, we conducted a PubMed search for articles published between January 1, 2011, and December 31, 2021, using the search terms *artificial intelligence* AND *skin cancer*. This search resulted in 809 articles which were screened for relevance. We also included the results of one prospective validation study for which we had conducted the analyses ourselves, but which had not yet been published at that time. Several commercial companies have implemented such algorithms in smartphone-based mobile health (mHealth) applications, making them available to the general public. A systematic review and meta-analysis reported that AI-based apps assessing skin cancer risk from macroscopic images achieved varying sensitivity and specificity, depending on the algorithm and the type of skin cancers detected. Four studies had prospectively validated the specific mHealth app investigated in this study against histopathology, with reported sensitivities ranging from 57% to 87% and specificities from 27% to 83%. Despite promising indications, no randomised controlled trials have yet evaluated the effectiveness of AI-based mHealth apps for skin cancer screening in the general population.

**Added value of this study:** The SPOT study is, to our knowledge, the first randomised controlled trial to investigate how implementing an AI-based skin cancer risk detection app in the general population affects skin cancer detection and healthcare use for benign skin lesions. We found a modestly higher skin cancer incidence amongst those who were offered to use the app, though this difference was not statistically significant. However we also found those who were offered to use the app had a significantly larger increase in healthcare visits and procedures for benign skin lesions. Suggesting that implementation in the general population may involve a possible trade-off between increased skin cancer detection and unnecessary care due to overdiagnosis.

**Implications of all the available evidence:** Even though research in a sterile setting shows potential for implementation of AI-based mHealth apps, the results from this study suggests that nationwide implementation of an mHealth with its current accuracy is not the most optimal strategy. Targeted implementation in higher-risk populations may offer a more favourable balance between benefits and harms.

## INTRODUCTION

Artificial intelligence (AI) plays an increasingly prominent role in medicine. Commercial companies have already introduced multiple direct-to-consumer AI algorithms to the general public through mobile health (mHealth) smartphone apps, some of which enable consumers to self-assess suspicious skin lesions.^1^ ^2^

Hypothesized benefits of offering such apps to the general population include increased detection of skin cancer and detection at an earlier stage. This is especially relevant for melanoma, where late-stage detection negatively affects survival.^3^ However, introducing these apps to the general population resembles national cancer screening programs, with similar issues like diagnostic delay due to false negative assessments and overdiagnosis due to false positives.^4^ Moreover, conventional population-based skin cancer screening through full-body skin examinations by a healthcare provider is generally not recommended given limited evidence of impact on mortality and cost-effectiveness.^5^ It is unclear if technological innovations such as these mHealth apps might change this paradigm.

While regulatory agencies strive to enforce devices to uphold a certain safety standard before they enter the market, experts agree that impact on care should also be monitored and evaluated after implementation.^6^ So far, only one Dutch population-based study based on insurance claims suggested that implementation of an mHealth app led to an increase in claims for cutaneous malignancies, but also increased care consumption for benign skin lesions.^7^ Because of its retrospective design it was unclear how much of this effect was due to app usage. Furthermore the study did not differentiate between skin cancer types and stage.

To address these issues, we conducted a randomized controlled trial (RCT) to evaluate the effect of implementation of an AI-based mHealth app in the general population on the types and stage of cutaneous (pre)malignancies detected as well as the healthcare consumption for benign skin lesions.

## METHODS

### Study design and participants

The SmartPhone applicatiOn for skin cancer risk assessmenT (SPOT) study is an investigator-initiated and -designed, unblinded, RCT. The Medical Ethics Committee of the Erasmus University Medical Center reviewed the trial and exempted it from the need for further ethical approval (MEC-2021-0180) as it did not fall under the scope of the Medical Research Involving Human Subjects Act (WMO) since the investigated mHealth app is already available to the general population. All participants gave informed consent for participation.

The study protocol was preregistered, and the trial was reported according to the CONSORT-AI checklist (Supplemental File).^8^ ^9^

240,819 randomly selected adult costumers of DSW (a not-for-profit health insurer) living in-and around Rotterdam the Netherlands were invited once through an email to participate in this trial between August and December 2021. DSW is the fifth largest health insurer in the Netherlands with a total of ∼675000 insurees. People were eligible for the study if they were over 18 years old and still insured with the participating health insurer. Exclusion criteria were withdrawal of consent, inability to link the data, and not having an email address or internet connection.

### Randomization and masking

Participants were randomized in the intervention or control group through a computer generated number sequence in a ratio of 3:2. We chose this ratio anticipating, based on a previous study, that not all individuals in the intervention group would use the app. ^7^ Randomization was not stratified. Since this was an open-label study, no masking was used.

### Procedures

The intervention group was offered free access to an AI-based mHealth app (SkinVision® SVS6.0.9, Amsterdam), available for iOS and Android smartphones, to assess self-identified suspicious skin lesions for an initial period of 12 months, which was later extended to 24 months. The app is clinically validated with a sensitivity of 87-95% and specificity of 70-78%.^10^ ^11^ As a build-in quality check, photos in the app can only be made if a lesion is centered, not covered (e.g., by hair), and the image is sharp. Each picture is assessed by a convolutional neural network (CNN), which generates a probability score for skin cancer risk. Based on the probability the user receives an automated message to either visit a doctor or to not worry about their lesion. Additionally, as per standard service of the app, all photos were assessed by a panel of three teledermatologists within 48 hours who could upgrade or downgrade the assessment if needed or ask the user to take a new photo if the picture quality was too low. The control group was not given free access to the app and instead followed care-as-usual, i.e. consulting their general practitioner (GP) and/or dermatologist if they worried about a lesion. A year after enrolment the controls were also granted 12 months of free access to the app (Figure S1). Participants from both groups were passively followed up beyond the initial 24-month period, with data collection ongoing at the time of analysis. The current analysis includes participants with up to 24 months of follow-up.

All participants completed an online questionnaire including items as self-reported skin type, educational level and ethnicity. Ethnicity and educational level were categorized according to national guidelines.^12^ ^13^ Participants age, sex, number of unique medications on ATC 5th level^14^, and healthcare claims were provided directly by the health insurer. Costs corresponding with the claims were computed based on fixed prices found in the public registry OpenDIS data.^15^ Cost for using the mHealth app were based on an annual business-to-business fee of €20.00 for all participants who used the app (i.e. made at least one assessment). The health insurer provided aggregated summary statistics of those who declined to participate in the study or did not respond to the study invitation (the non-responders).

Histopathology reports of cutaneous biopsies and excisions were collected from the Dutch Nationwide Pathology Databank (Palga).^16^ Re-excisions of lesions that were already present prior to the start of the study were not counted as new detected lesions. A history of skin cancer within the study population was defined as having a histologically verified malignant cutaneous tumour recorded in Palga within 10 years prior to the start of the study. For the non-responders, skin cancer history this was defined as presence of a health insurance claim for a malignant skin lesion within 5 years prior to this study. All data were linked through a unique identifier. Histopathology reports were pseudonymized and linked via a trusted third party (ZorgTTP) to ensure adherence to European privacy guidelines (Figure S2). Data on app usage (e.g., downloads, (un)successful number of images taken) was provided by SkinVision®.

### Outcomes

The primary outcome was the difference in healthcare consumption for (pre)malignant and benign skin lesions between the intervention and control group after 12-months. Care consumption was assessed by (1) the difference in incidence of histologically confirmed malignant skin tumours, including melanoma, cutaneous squamous cell carcinoma (cSCC), and basal cell carcinoma (BCC); (2) the difference in incidence of claims for benign skin tumours, defined as benign skin tumours, verrucae, and nevi; and (3) the difference in incidence of claims for (pre)malignant skin tumours (Table S1). Secondary outcomes were differences in melanoma characteristics (Breslow thickness and T-stage) at 12 months follow-up, the number of surgical interventions at the GP (primary care) and in a hospital-based setting (secondary care) at 12 months follow-up, the overall use of dermatological care at 12 months follow-up and a cross-sectional cost-effectiveness analysis.

### Statistical Analysis

Sample sizes were estimated prior to the start of the study to be able to detect a statistically significant difference in proportions of at least 20% based on a χ^2^ test (two-sided alpha of 0.05, power 80%) (Figure S3). Dutch yearly skin cancer incidence rate was estimated to be 1/190. Those willing to participate were expected to have a 43% (1/133) to 71% (1/111) higher skin cancer risk, compared to the general population. Therefore we calculated sample sizes for a range of skin cancer incidences. The cooperating health insurer only enabled us to invite a maximum 240,819 randomly selected individuals in the region in and around Rotterdam, therefore this was the total population invited to participate.

Differences in participants characteristics were assessed using t-tests for continuous variables and χ^2^ for categorical variables. Some participants had missing data in their baseline characteristics, this was reported when applicable and participants were not excluded from analyses. The primary outcomes were tested using χ^2^ tests. For the secondary outcomes we tested differences in melanoma T-stage using Fisher’s exact test and differences in Breslow thickness using Wilcoxon’s rank test. Differences in number of surgical interventions at a primary and secondary care level and differences in proportion of participants with a new consultation at the dermatologist were compared using χ^2^ tests. Specific categories included in the analyses are described in further detail in Table S2. Additionally, differences in costs were compared based on dermatological claims data and the costs for using the mHealth app over a time horizon of 12 months using t-tests. As a sensitivity analysis, we investigated differences between app-users and non-app-users, regardless of their randomization, by looking at their baseline characteristics, incidence of tumours, and claims for benign and (pre)malignant skin lesions. These analyses were repeated for the second year of the study.

Analyses were performed in the intention-to-treat population, unless indicated otherwise. The Benjamini-Hochberg procedure was applied to control the false discovery rate. Adjusted p-values <0.05 were considered statistically significant. If no tests were statistically significant, no corrections were done. Analyses were performed using R (version 4.3.2).

## RESULTS

### Participant characteristics

Between August and December 2021, 240,819 adults were invited to participate in this study. Of those invited adults, 19,560 (8·1%) agreed to participate and 19,009 were included in the final analysis (Figure 1). Including 7,536 in the control group and 11,473 in the intervention group.

**Figure 1.**
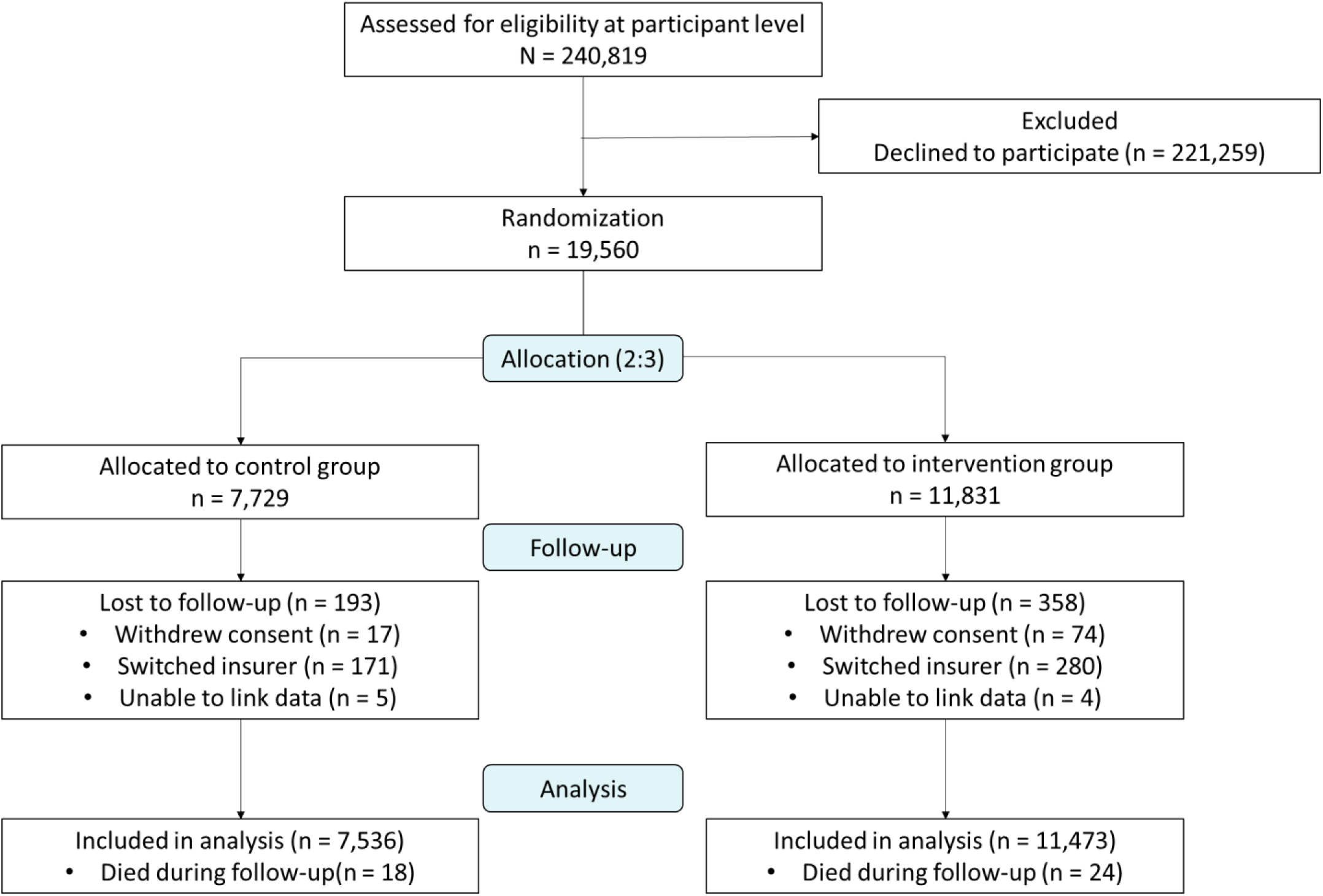
Enrolment, randomization and follow-up of participants.

The study population was older than the non-responders (mean 55·1 vs. 49·5 years; p<0·001), with a larger proportion of females (57·3% vs. 51·6%; p<0·001), and a larger proportion of prior skin cancer (5·1% vs. 1·5%; p<0·001) (Table S3). The intervention and control group were comparable in baseline characteristics (Table 1).

**Table 1.** Baseline characteristics of study participants.

|  | Control group<br>(N = 7,536 ) | Intervention group<br>(N = 11,473 ) |
| --- | --- | --- |
| Age, years | 55.1±14.2 | 55.1±14.2 |
| Sex |  |  |
| Male | 3,251 (43.1%) | 4,875 (42.5%) |
| Female | 4,285 (56.9%) | 6,598 (57.5%) |
| Self-reported skin type |  |  |
| Very white | 3,384 (44.9%) | 5,039 (43.9%) |
| White with a beige tone | 3,669 (48.7%) | 5,713 (49.8%) |
| Light brown | 433 (5.8%) | 650 (5.7%) |
| Dark brown | 50 (0.7%) | 71 (0.6%) |
| Ethnicity |  |  |
| Dutch | 6,924 (91.9%) | 10,564 (92.1%) |
| European other | 260 (3.5%) | 381 (3.3%) |
| Non-European | 334 (4.4%) | 503 (4.4%) |
| Missing | 18 (0.2%) | 25 (0.2%) |
| Education |  |  |
| Low | 1,288 (17.1%) | 2,018 (17.6%) |
| Middle | 2,568 (34.1%) | 4,074 (35.5%) |
| High | 3,490 (46.3%) | 5,122 (44.6%) |
| Missing | 190 (2.5%) | 259 (2.3%) |
| Keratinocytic skin cancer in medical history* | 328 (4.4%) | 504 (4.4%) |
| Melanoma in medical history* | 57 (0.8%) | 81 (0.7%) |
| Median number of medications† | 2 (1-5) | 3 (1-5) |

Data are mean (±SD), median (IQR) or n (%) Differences between groups were compared with t-tests for means and medians and $\chi^2$ for categorical variables. IQR = interquartile range.
\*Excluding metastases
† Median number of medications in the year before enrolment in the study, according to the ATC5th level

At 12 months after randomization 77·6% (n = 8,901) of the intervention group downloaded the app and 53·9% (n = 6,179) had one or more successful assessments from the algorithm (median of 2 assessments (IQR 1 - 5)). From the total of 24,666 pictures submitted via the app, 22,803 (92·4%) were assessed by the CNN of which 15·1% (n = 3,449) was rated high risk and 84·9% (n = 19,354) as low risk. The teledermatologists upgraded 0·7% (n = 171) of the algorithms assessments, downgraded 9·9% (n = 2,263), and 2·9% (n = 670) was deemed as unassessable. Despite randomization, 0·7% (n = 51) of the control group used the app on their own initiative. Between 12 and 24 months after randomization the active number of app-users in the intervention group decreased to 16·2% (n = 1,855) and 17·6% (n=1,329) of the control group became app-users (Figure S4).

### Incidence of (pre)malignant and benign skin lesions

At 12 months, the overall incidence of histologically verified malignant skin tumour was higher in the intervention group (2·7% (n = 305) vs. 2·3% (n = 171), risk difference 0·4% 95% confidence interval (CI) -0·1 to 0·9), although not statistically significant (p = 0·10). Interpretation of this finding is limited by the study’s statistical power No notable differences were observed for the skin cancer subtypes (Table 2). The histologically confirmed melanomas had a similar Breslow thickness and T-stage distribution. The incidence of claims for a benign skin lesion in a hospital-based dermatology setting was significantly higher in the intervention (3·9%, n = 443) than the control group (2·6%, n = 198, risk difference 1·3% 95% CI 0·7 to 1·7, p<0·001) (Table 3). In line with the pathology reports, the incidence of claims for a (pre)malignant skin lesion in a hospital-based dermatology setting was similar between both groups. Notably, the incidence of claims was 3·4-fold lower among the non-responders for benign skin lesions (1·0%, n = 2,034, risk difference 2·4 95% CI 2·1 to 2·7, p<0·001) and 2·8-fold lower for (pre)malignant skin lesions (2·4% n = 4,981, risk difference 4·3, 95% CI 3·9 to 4·6, p<0·001).

**Table 2.** Number of histologically confirmed melanomas, cutaneous squamous cell carcinomas and basal cell carcinomas within the control and intervention group at 12 months follow-up.

|  | Control group<br>(N = 7,536) | Intervention group<br>(N = 11,473) | Risk Difference<br>(95% CI) | p-value |
| --- | --- | --- | --- | --- |
| Melanoma | 23 | 30 |  |  |
| Patients with a melanoma | 23 (0.31%) | 30 (0.26%) | -0.05% (-0.21 - 0.12) | 0.68 |
| Median Breslow thickness (IQR) - mm | 0.7 [0.4-0.9] | 0.5 [0.3-0.7] |  | 0.59 |
| Melanoma TNM stage* |  |  |  | 0.38 |
| pTis | 11 (47.8%) | 8 (26.7%) |  |  |
| pT1a & pT1b | 10 (43.5%) | 17 (56.7%) |  |  |
| pT2a & pT2b | 2 (8.7%) | 2 (6.7%) |  |  |
| pT3a & pT3b | 0 (0%) | 1 (3.3%) |  |  |
| pT4a & pT4b | 0 (0%) | 2 (6.7%) |  |  |
| cSCC – no. | 22 | 61 |  |  |
| Patients with a cSCC – no. (%) | 22 (0.29%) | 55 (0.48%) | 0.19% (0 - 0.37) | 0.06 |
| BCC – no. | 196 | 292 |  |  |
| Patients with a BCC– no. (%) | 135 (1.79%) | 228 (1.99%) | 0.2% (-0.21 - 0.6) | 0.36 |
| Total – no. † | 244 | 383 |  |  |
| Patients with any type of skin cancer – no. (%)† | 171 (2.27%) | 305 (2.66%) | 0.39% (-0.07 - 0.85) | 0.10 |

Data are n or n (%) unless otherwise indicated. Differences between groups were compared Wilcoxon's rank sum test for medians, $\chi^2$ for categorical variables containing at least 10 observations within a cell and Fisher's exact test for categorical variables containing less than 10 observations within a cell. IQR = interquartile range, mm = millimetre, CI = confidence interval, cSCC = cutaneous squamous cell carcinoma, BCC = Basal cell carcinoma. Percentages may not sum to 100 because of rounding.
\* Staging reported according to the American Joint Committee on Cancer eighth edition (AJCC8).
† Only including melanoma, cSCC and BCC.

**Table 3.** Proportion of participants with a claim for a benign and/or (pre)malignant skin lesion within the control and intervention group, and non-responders at 12 months follow-up.

|  | Control<br>group<br>(N = 7,536) | Intervention<br>group<br>(N = 11,473) | Risk Difference<br>(95% CI) | Adjusted<br>p-value* | Non-responders<br>(N = 207,090) | Risk<br>Difference<br>(95% CI) | Adjusted<br>p-value† |
| --- | --- | --- | --- | --- | --- | --- | --- |
| **Benign skin tumours and nevi** |  |  |  |  |  |  |  |
| Benign skin lesions, dermatology-based claims | 198 (2.6%) | 443 (3.9%) | 1.3% (0.7 - 1.7) | < 0.001 | 2,034 (1.0%) | 2.4% (2.1 - 2.7) | < 0.001 |
| Benign skin lesions, claims from all relevant specialties | 235 (3.1%) | 506 (4.4%) | 1.3% (0.7 - 1.8) | < 0.001 | 2,806 (1.4%) | 2.5% (2.3 - 2.8) | < 0.001 |
| **Premalignant and malignant skin lesions** |  |  |  |  |  |  |  |
| (Pre)malignant skin lesions, dermatology-based claims | 478 (6.3%) | 791 (6.9%) | 0.6% (-0.2 - 1.3) | 0.23 | 4,981(2.4%) | 4.3% (3.9 - 4.6) | < 0.001 |
| Of which premalignant only | 333 (4.4%) | 557 (4.9%) | 0.5% (-0.2 - 1.1) | 0.23 | - | - | - |
| Of which malignant only | 153 (2.0%) | 244 (2.1%) | 0.1% (-0.3 - 0.5) | 0.69 | - | - | - |
| (Pre)malignant skin lesions, claims from all relevant specialties | 494 (6.6%) | 810 (7.1%) | 0.5% (-0.2 - 1.2) | 0.23 | 5,102 (2.5%) | 4.4% (4 - 4.8) | < 0.001 |

Data are n or n (%) unless otherwise indicated. CI = Confidence interval.
\* P-values reflect the difference between the control and intervention group calculated using a $\chi^2$ test.
† P-values reflect the difference between the study population (control and intervention group combined) and non-responders calculated using a $\chi^2$ test.

### Procedures and consultations

In primary care, a larger proportion of participants from the intervention group underwent a biopsy or excision for benign skin tumours and nevi (3·2% (n = 367) vs. 2·6% (n = 197), risk difference 0·6%, 95% CI 0·1 to 1·1 p = 0·04) and (pre)malignant skin lesions (1·7% (n = 194) vs 1·1% (n = 85), risk difference 0.6%, 95% CI 0·2 to 0·9, p = 0·005) (Table 4). In a hospital-based dermatology setting, participants from the intervention group had more consultations (2·7% (n = 312) vs 1·9% (n = 145), risk difference 0.8%, 95% CI 0·4 to 1·2 p = 0·002) and more biopsies and excisions of benign skin tumours and nevi (2·2% (n = 253) vs 1·5% (n = 113), risk difference 0·7%, 95% CI 0·3 to 1·1 p = 0·002), but not for (pre)malignancies. A detailed breakdown of the procedures for benign skin tumours and naevi can be found in Figure S5.

**Table 4.** Number of biopsies, excisions, and dermatological consultations within the intervention and control group at 12 months follow-up.

|  | Control group<br>(N = 7,536) | Intervention group<br>(N = 11,473) | Risk Difference<br>(95% CI) | Adjusted p-value |
| --- | --- | --- | --- | --- |
| **Benign skin tumours and nevi** |  |  |  |  |
| Biopsy or excision in primary care* |  |  |  |  |
| Total number of procedures | 230 | 415 |  |  |
| Patients with a procedure | 197 (2.61%) | 367 (3.2%) | 0.59% (0.09 - 1.08) | 0.04 |
| Biopsy or excision in hospital setting† |  |  |  |  |
| Total number of procedures | 136 | 311 |  |  |
| Patients with a procedure | 113 (1.5%) | 253 (2.21%) | 0.71% (0.31 - 1.1) | 0.002 |
| New dermatological consultations |  |  |  |  |
| Total number of consultations | 165 | 346 |  |  |
| Patients with a consultations | 145 (1.92%) | 312 (2.72%) | 0.8% (0.35 - 1.24) | 0.002 |
| **Premalignant and malignant lesions** |  |  |  |  |
| Biopsy or excision in primary care* |  |  |  |  |
| Total number of procedures | 122 | 259 |  |  |
| Patients with a procedure | 85 (1.13%) | 194 (1.69%) | 0.56% (0.22 - 0.91) | 0.005 |
| Biopsy or excision in hospital setting† |  |  |  |  |
| Total number of procedures | 313 | 512 |  |  |
| Patients with a procedure | 186 (2.47%) | 316 (2.75%) | 0.28% (-0.19 - 0.76) | 0.29 |
| New dermatological consultations |  |  |  |  |
| Total number of consultations | 443 | 757 |  |  |
| Patients with a consultations | 351 (4.66%) | 578 (5.04%) | 0.38% (-0.25 - 1.01) | 0.29 |
| Mohs surgery† |  |  |  |  |
| Total number of procedures | 36 | 47 |  |  |
| Patients with a procedure | 35 (0.46%) | 44 (0.38%) | -0.08% (-0.28 - 0.12) | 0.46 |

Data are n or n (%) unless otherwise indicated. P-values reflect the difference in proportion of patients with a procedure per category calculated using a $\chi^2$ test.
\*Biopsies or excisions performed in a primary care setting are done by a general practitioner.
†The described excisions in a hospital setting are usually done by a dermatologist or dermatology resident.

### Healthcare costs

Since no statistically significant differences were found in detected skin cancers between the intervention and control group, a formal cost-effectiveness analysis was not performed. The overall mean costs of care were 34·4% higher in the intervention group (€63 (95% CI 58 to 67); vs. €47 (41 to 52); p<0·001) (Table S4). The increased costs were mainly related to the additional cost for using the app and a difference in the mean costs related to claims for nevi (€7 (6 to 8); vs. €4 (CI 3 to 5); p<0·001). The mean and median costs per claim subtype, for participants who had at least one claim within a category, were similar between both groups (Figure S6 and Table S5).

### App-users versus non app-users

App-users were younger, more often female, with a Dutch ethnicity, had a lighter skin type, and received a higher level of education in both the first and second year of the study (Table S6 and S7). More skin cancer, primarily BCCs, was detected amongst app-users in the first (3·0% (n = 184) vs. 2·3% (n = 292), risk difference 0·7% 95% CI 0·2 to 1·2, p = 0·03) and the second year (2·8% (n = 90) vs. 2·0% (n = 320), risk difference 0·8% 95% CI 0·2 to 1·4, p= 0·01) (Table S8 and S9). This difference in BCCs was not significant after multiple testing correction at 12 months, but remained significant at 24 months (2·4% (n = 75) vs. 1·5% (n = 236), risk difference 0·9% 95% CI 0·3 to 1·4, p = 0·002). The increased skin cancer detection was again accompanied by a larger proportion of app-users with claims for benign skin tumours or nevi at both 12 months follow-up (4·9% (n = 304) vs. 2·6% (n = 337), risk difference 2·3% 95% CI 1·6 to 2·9, p<0·001) and 24 months follow-up (4·8% (n = 154) vs 2·3% (n = 369), risk difference 2·5% 95% CI 1·7 to 3·3, p < 0·001) (Table S10 and S11).

## DISCUSSION

In this RCT, there was a higher but non-statistically significant incidence of skin cancer in participants with access to an AI-based mHealth app to assess suspicious skin lesions compared to controls randomized to care-as-usual. However, participants who were offered access to an mHealth app had significantly more dermatological medical procedures for benign skin lesions than those who were not resulting in higher direct healthcare costs.

Although the intention-to-treat analyses did not reveal a statistically significant beneficial impact of AI-assisted mHealth apps, the results provide relevant insights into the short-term effects of screening-like interventions for skin cancer in the general population. A previous non-randomized retrospective claim-based study demonstrated that app-users had more claims for cutaneous malignancies than non-users.^7^ Comparing app-users vs non app-users in this RCT suggests that most of these additional skin cancers were BCCs. However, these effects might be partially explained by selection bias, reflecting inherent characteristics of participants. Supporting this, the overall skin cancer incidence in participants of this RCT was much higher than in non-responders and in the general population^17^, but comparable between the intervention and control group. Arguing that those who were interested in using this app represent a different type of population than the general population. It could be that participants represent a group who are more worried about their skin or think they have a higher risk of developing skin cancer. In other words, the high skin cancer incidence among the study population suggests that participants likely had prevalent skin cancers and that the study invitation induced an initial increase in skin cancer detection in a population already primed and more aware of their suspicious skin lesions. Such an initial increase in incidence of malignancies is often seen in screening studies.^18^ ^19^ However, it might have been more substantial because skin cancer is externally visible and can be self-detected, unlike internal cancers, which typically require medical imaging or diagnostic tests. Therefore, it could be argued that the high skin cancer incidence in this RCT was driven by the act of creating awareness amongst a high-risk population, rather than app usage itself.

The introduction of new screening-like interventions typically leads to an increase in false positives. This also occurred within the intervention group of this RCT, especially among participants who used the app, resulting in increased care consumption and higher healthcare costs for benign skin lesions. The false positives were noted among GPs and dermatologists performing additional procedures for potentially suspicious lesions that turned out to be benign. There is currently no evidence to justify full-body skin cancer screening in the general population and it remains questionable whether AI-based mHealth apps might change this based on this study.^5^ Implementation in populations with a higher skin cancer prevalence might be more beneficial. For example, AI-based mHealth apps could replace low-value follow-up consultations^20^, or serve as triage tools or diagnostic aids in primary care to streamline the care pathway of people with suspicious skin lesions.^21^ ^22^ These approaches are more in line with and might enhance lesion-directed screening, which is found to be equally effective and possibly even more cost-effective for skin cancer than full-body skin examinations.^23^

Despite free access to the mHealth app, the uptake was relatively low, respectively 53·9% in the first year and only 16·7% in the second. As a comparison, the uptake for national breast, cervical and colorectal cancer screening programs in the Netherlands varies from 65% to 75%.^24^ That the uptake was already lower within a study of probably more motivated people could partly be explained by a lack of concern about skin cancer, usability issues or distrust in mHealth apps and AI in the general population.^25^ ^26^ Looking into who used the app, we found that app-users were significantly younger and more often had a lighter self-reported skin type. The younger age among app-users might partially be explained by technological literacy or different attitudes towards usage of technology for health issues across different age groups.^27^ ^28^ Since skin cancer incidence varies across populations with different skin types, this might explain the larger proportion of participants with lighter skin types using the app.^29^ However, it should be safeguarded that this difference was not due to usability issues, as algorithms have been shown to be at risk of bias in the assessment of lesions on darker pigmented skin.^30^ Additional attention should be given to improve implementation and user engagement.

To our knowledge, this is the first RCT that investigates the direct impact of an AI-based mHealth app on skin cancer detection and care consumption in the general population.

Moreover, it is to our knowledge one of the largest randomized skin cancer screening studies conducted to date and the randomised design allows for evaluation of causality. However, there were some limitations to this trial. First, even though the skin cancer incidence was much higher than previously anticipated, the study was underpowered for the first primary outcome, namely the difference is histologically confirmed malignant skin tumours (N= 19,009, incidence 1/39, delta 17%, power 52%). We conducted a post-hoc power analysis to evaluate how big the sample size should have been to detect a significant difference of 20% with this incidence given the 3:2 randomization ratio and found this should have been 38,551 (Figure S3). However, the study was powered for the other primary outcomes. Second, follow-up time for the primary outcomes was limited to one year. App usage could change over a longer period of time and to evaluate impact on mortality the follow-up should be longer. Third, while the RCT limits the risk of selection bias and confounding through randomization, unintended crossover of participants and non-compliance may still affect the results. We conducted subgroup analyses to explore this potential impact, which suggested a higher skin cancer incidence amongst app-users. However, this approach may introduce selection bias, limiting causal interpretation. Fourth, it could be that only highly motivated individuals or those with a specific focus on their skin cancer risk participated, limiting generalisability. However, the high skin cancer incidence observed among participants compared to non-responders suggests that people were quite effective at self-selecting into the study based on their perceived high risk.

In conclusion, in this trial we observed a modest, but not statistically significant, higher skin cancer incidence in those with access to an AI-based app compared to care-as-usual after 12 months. This was accompanied by a larger statistically significant difference in care consumption for benign skin lesions. During an extended follow-up of 24 months, significantly more BCCs were found amongst app-users in a non-randomized setting.

Therefore, we recommend conducting population-based studies with longer follow-up periods or focusing on targeted high-risk populations.

## Contributors

ASG drafted the manuscript, assisted by MW and TN. All authors reviewed the manuscript, provided critical feedback and approved it. ASG, TS, CUG, MW, TN were responsible for the design of the trial. ASG was responsible for trial conduct and logistics. ASG was responsible for database design and management. ASG was responsible for the analyses, assisted by TS, CUG, MW and TN. All authors contributed to the interpretation of the data. All authors had full access to all the data in the study and had final responsibility for the decision to submit for publication. ASG directly accessed and verified the underlying data reported in this manuscript. All authors read and approved the final manuscript. MW is guarantor. The corresponding author attests that all listed authors meet authorship criteria and that no others meeting the criteria have been omitted.

## Data sharing

The data collected for this study were combined across multiple healthcare systems through mutual Data Transfer Agreement and under approval of an Institutional Review Board.

Therefore, data will not be made publicly available. Study protocol, statistical analysis plan and analytic code can be shared on request for academic purposes. Proposals should be directed to.

## Data Availability

The data collected for this study were combined across multiple healthcare systems through mutual Data Transfer Agreement and under approval of an Institutional Review Board. Therefore, data will not be made publicly available. Study protocol, statistical analysis plan and analytic code can be shared on request for academic purposes. Proposals should be directed to.

## Acknowledgments

We thank the Erasmus MC Datacapture team in their support for building the registration platform and questionnaire.

