## Supplemental Material for "The impact of a SmartPhone applicatiOn for skin cancer risk assessmenT on the healthcare system (SPOT-study): A randomized controlled trial"

#### Supplementary Appendix

### Supplemental Figures

**Figure S1.** Graphical overview of the trial design.

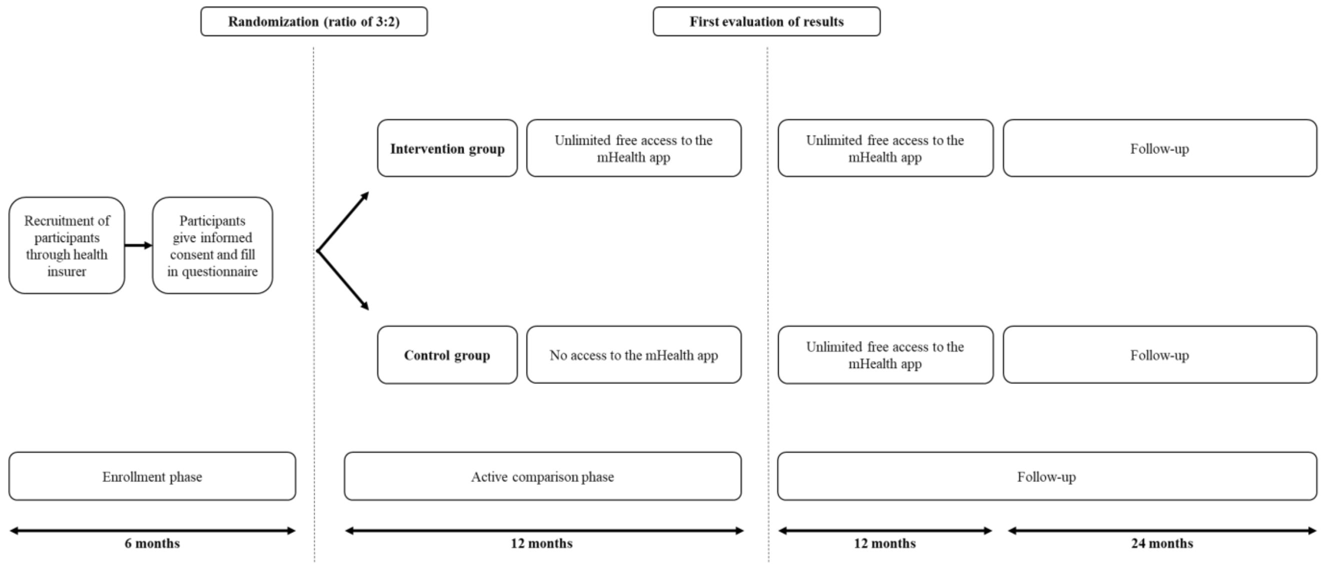

**Figure S2.** Schematic overview of data linkage and collection

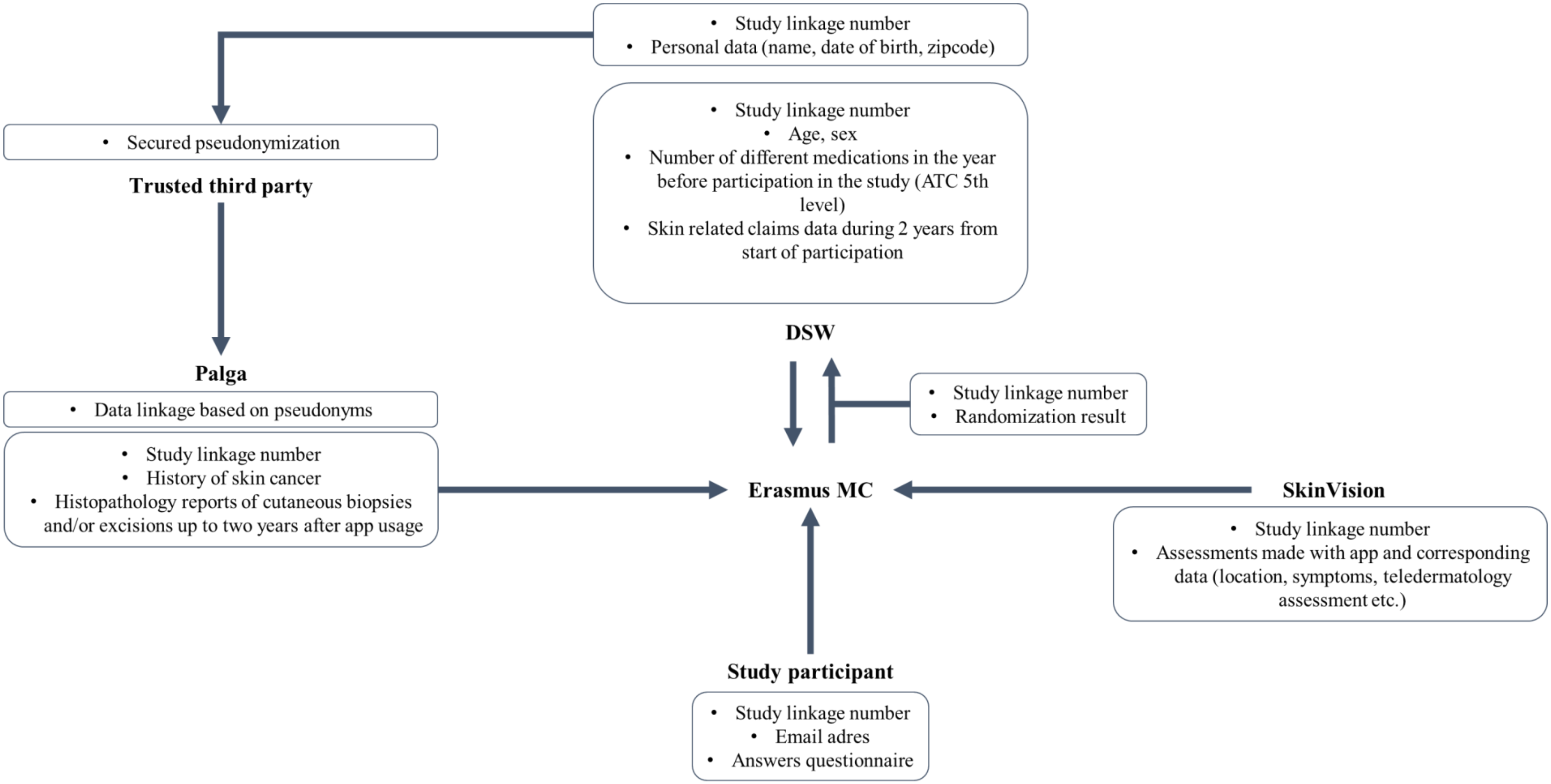

**Figure S3.** Figure of sample size calculations stratified for multiple incidences of skin cancer and post-hoc sample size calculation with skin cancer incidence found in the study.

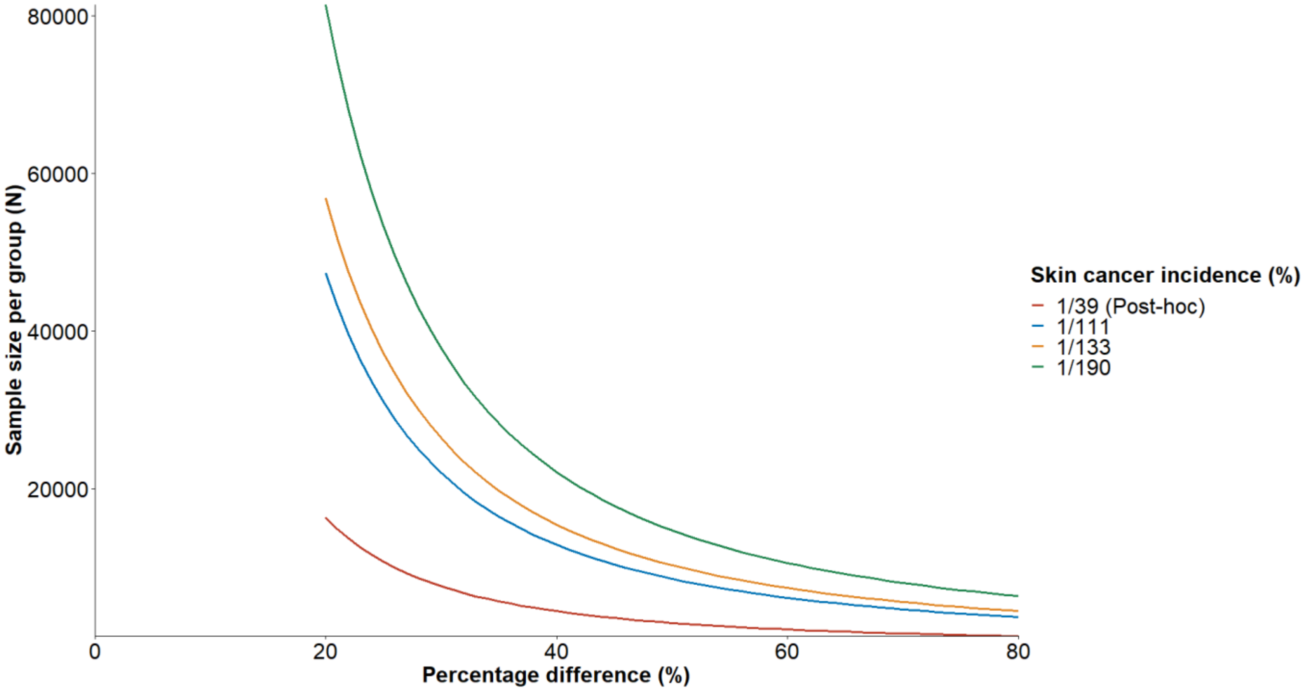

**Figure S4** Sankey plot displaying the flow of participants becoming app-users during the first and second study year.

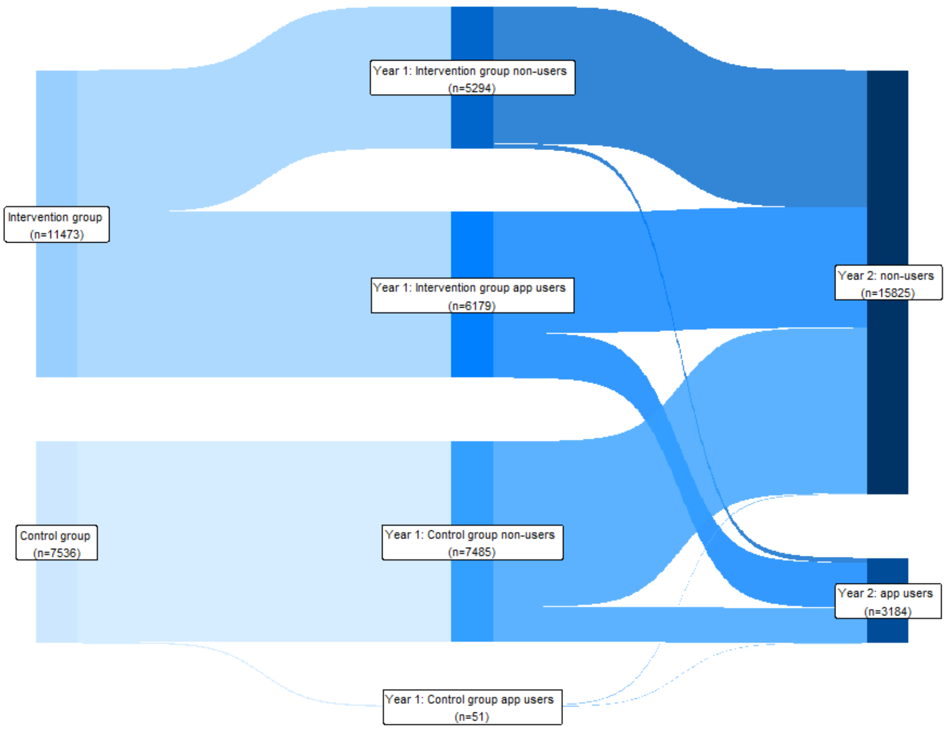

**Figure S5** Detailed flowchart describing the procedures performed for benign skin lesions in a hospital based setting in the first year after randomization.

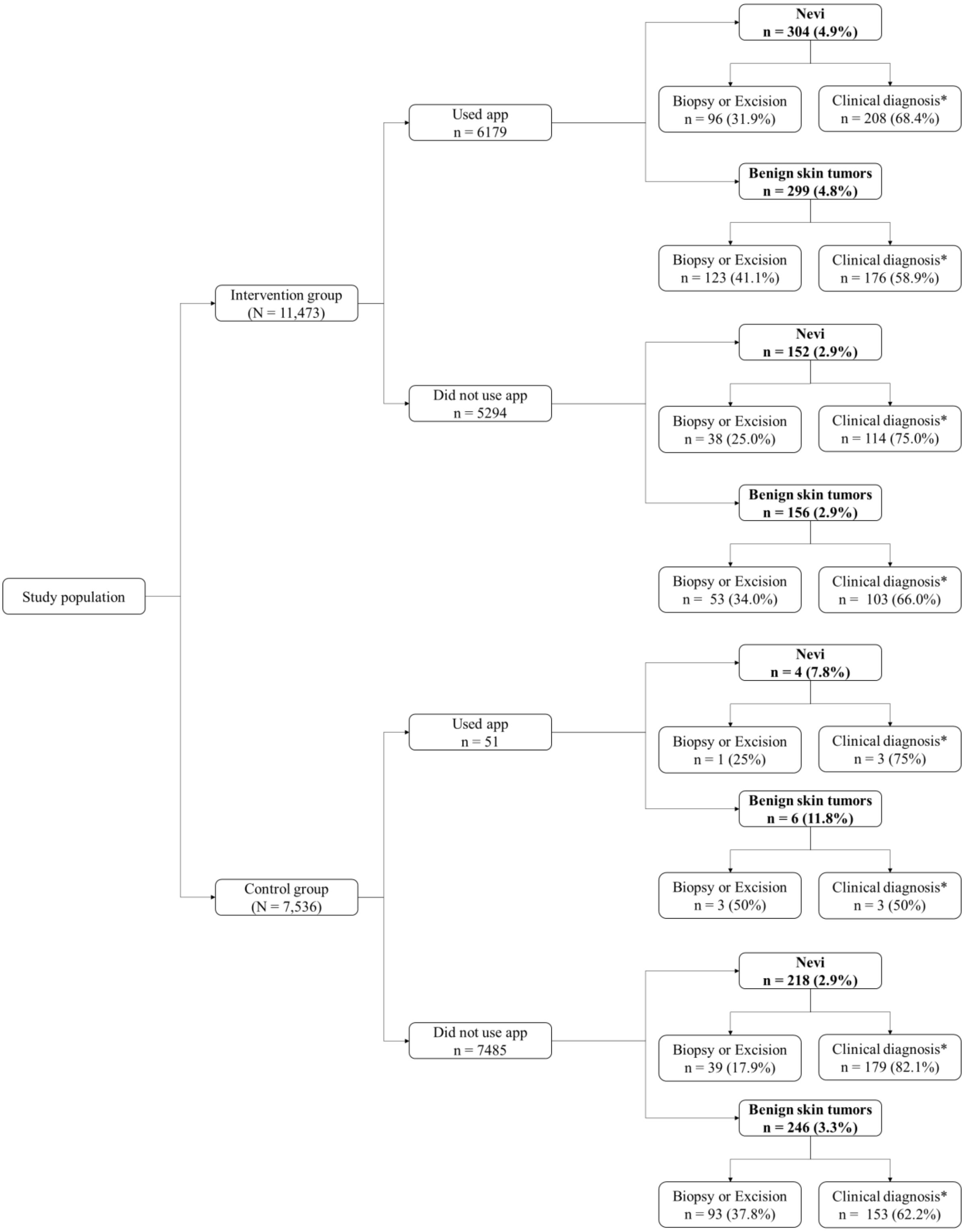

*Clinical diagnosis represents patients with a clinical diagnosis, not the absolute number of lesions assessed.

Biopsies and/or excisions represent the absolute number of lesions.

**Figure S6**. Boxplot of the distribution of costs per made per person, per category of claims data for participants of the intervention and control group who had at least one claim within the category in the first year after randomization.

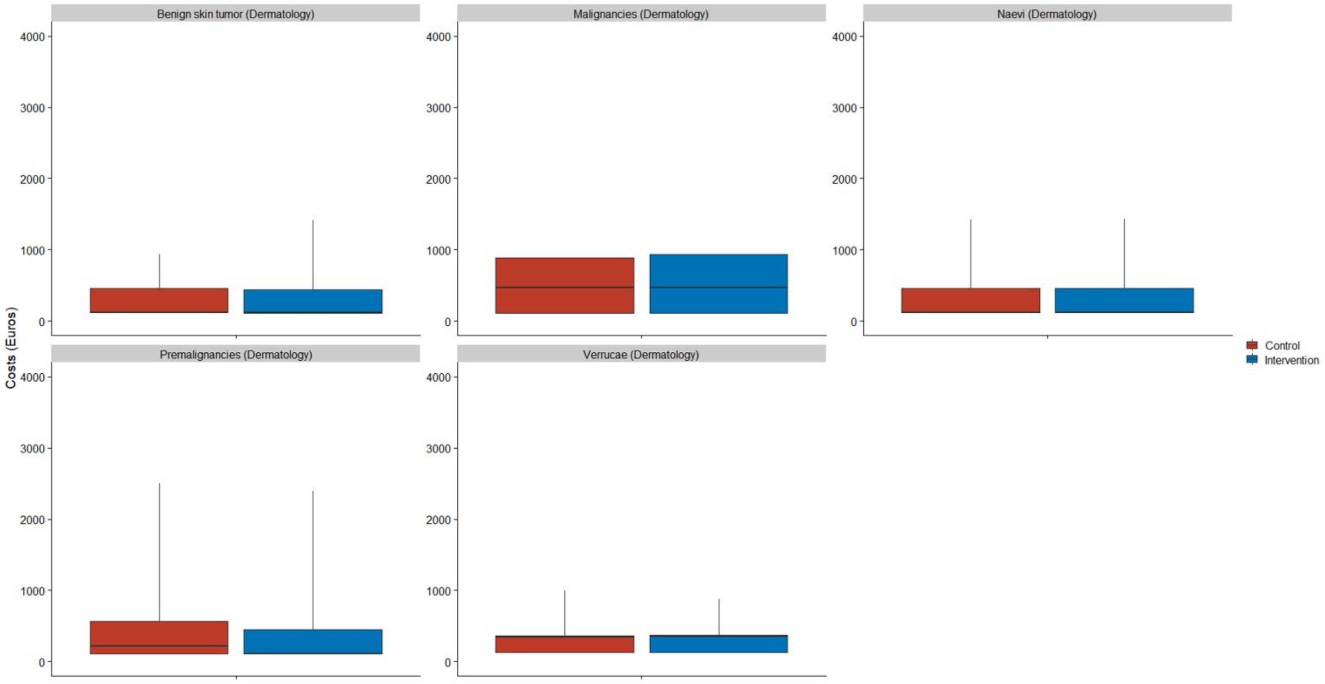

### Supplemental Tables

**Table S1.** Specific subtypes of claims (DBC codes) included in the analysis of the primary outcome.

| **Specialty** | **Specialty code** | **Benign skin lesions – Included diagnosis codes** |
|---|---|---|
| Dermatology | 310 | 3 - Benign tumours |
| | 310 | 15 - Nevi |
| | 310 | 26 - Verrucae |
| Surgery | 303 | 171 – Nevus, small lipoma, atheroma |
| ENT | 302 | 87 - Infection/benign skin tumour head/neck |
| Plastic surgery | 304 | 508 - Excision 1-3 benign tumours/ nevi not in FG, scar correction not FG with transposition or transplant < 1% |
| | 304 | 510 - Excision 1-3 benign. tumours / nevi w.o. in FG or 4-10 not in FG |
| | 304 | 512- Excision benign tumour / PW and transposition or transplant 1-3% not in FG, or, 4-10 nevi w.o. in FG |
| | 304 | 514 - Excision benign tumors / PW for which transposition or transplant >3% not in FG, more than 10 benign tumours / nevi |
| **Specialty** | **Specialty code** | **(Pre)malignant skin lesions - Included diagnosis codes** |
| Dermatology | 310 | 14 – Malignant dermatosis |
| | 310 | 17 - Premalignant dermatosis |
| Internal medicine | 313 | 842- Malignancy of the skin/melanoma |
| Surgery | 303 | 350 – Malignant melanoma |
| ENT | 302 | 88- Malignant skin tumours head/neck |
| Plastic surgery | 304 | 509 - Malignant tumour not in FG |
| | 304 | 511 - Malignant tumour in FG for which transposition or transplant. <1% |
| | 304 | 513 - Excision malignant tumour/PW requiring transposition or transplant in FG 1-3% or not in FG > 3%, 2-5 malignant tumours |

**Table S2.** Specific subtypes of claims (DBC codes) and procedure codes included to test differences in dermatological care.

| | Data source | Diagnosis code | Procedure code |
|---|---|---|---|
| **Benign skin tumours and nevi** | | | |
| Biopsy or excision in primary care | Pathology data | NA | NA |
| Excision in hospital setting | Pathology data and insurance data | 3, 15, 26 | 029899007, 029899004, 029899008, 029899005, 029499021, 029499016, 029499022, 029499017, 029499003, 0129999037, 0129999093, 0129999036 |
| New dermatological consultation | Insurance data | 3, 15, 26 | 029499039, 029499034, 029899013, 029899011, 011101010 , 011101008 |
| **Premalignant and malignant lesions** | | | |
| Biopsy or excision in primary care | Pathology data | NA | NA |
| Excision in hospital setting | Pathology data and insurance data | 14, 17 | 029899007, 029899004, 029899008, 029899005, 029499021, 029499016, 029499022, 029499017, 029499003, 0129999037, 0129999093, 0129999036 |
| New dermatological consultation | Insurance data | 14, 17 | 029499039, 029499034, 029899013, 029899011 |
| Mohs surgery | Insurance data | 14, 17 | 29499002 |

NA = not applicable.

**Table S3.** Baseline characteristics of study participants and non-responders.

| | **Study participants** (N = 19,009) | **Non-responders** (N = 220,975) | **p-value** |
|---|---|---|---|
| Age, years | 55·1±14·2 | 49·5±17·5 | < 0·001 |
| Sex | | | < 0·001 |
| Male | 8,126 (42·7%) | 106,974 (48·4%) | |
| Female | 10,883 (57·3%) | 114,001 (51·6%) | |
| Cutaneous malignancy in medical history | 970 (5·1%) | 3,278 (1·5%) | < 0·001 |
| Missing | - | 35,090 (15·9%) | |
| Median number of medications* | 3 (1 - 5) | 3 (2 - 6) | -† |

Data are mean (±SD), median (IQR) or n (%). Differences between groups were compared with t-tests for means and $\chi2$ for categorical variables. IQR = interquartile range.
* Median number of medications in the year before enrolment in the study, according to the ATC5th level.
† Since only aggregated summary statistics were available for the median number of medications for the non-responders a p-value could not be computed.

**Table S4**. Comparison of the mean dermatological health care related costs between the control group (n = 7,536) and the intervention group (n = 11,473) at 12 months follow-up.

| | Control group, mean (95% CI) (€) (n = 7,536) | Intervention group, mean (95% CI) (€) (n = 11,473) | Adjusted p-value |
|---|---|---|---|
| Cost mHealth app | 0 | 10·77 (10·59 - 10·95) | NA |
| Benign skin tumours | 2·43 (1·72 - 3·15) | 2·99 (2·33 - 3·65) | 0·40 |
| Nevi | 4·1 (3·07 - 5·12) | 7·2 (6·08 - 8·31) | < 0·001 |
| Verrucae | 0·68 (0·29 - 1·07) | 0·82 (0·48 - 1·16) | 0·59 |
| Premalignant skin lesions | 7·68 (5·83 - 9·53) | 6·32 (5·16 - 7·49) | 0·40 |
| Malignant skin lesions | 31·78 (26·52 - 37·03) | 34·7 (30·42 - 38·98) | 0·48 |
| Total | 46·71 (40·99 - 52·43) | 62·8 (58·17 - 67·42) | < 0·001 |

Costs are mean costs per person for each subcategory based on the number and types of claims multiplied with costs derived from the open DIS data. Total costs are the mean cost per person for all the categories combined, including cost for app usage. The adjusted p-values were determined by comparing means using an unpaired two-sided t-test. CI = confidence interval, NA = not applicable.

**Table S5**. Costs per category of claims data for participants of the intervention and control group who had at least one claim within the category at 12 months follow-up.

| | **Control group** | **Intervention group** | **Adjusted p-value** |
|---|---|---|---|
| Benign skin tumours | | | |
| Number of persons | 72 | 140 | |
| Mean € (95% CI) | 254·79 (207·54 - 302·04) | 244·86 (208·36 - 281·35) | 0·96 |
| Nevi | | | |
| Number of persons | 112 | 274 | |
| Mean € (95% CI) | 275·8 (228·29 - 323·31) | 301·33 (270·37 - 332·29) | 0·94 |
| Verrucae | | | |
| Number of persons | 17 | 31 | |
| Mean € (95% CI) | 301·47 (194·57 - 408·37) | 304·19 (235·54 - 372·85) | 0·96 |
| Premalignant skin lesions | | | |
| Number of persons | 153 | 244 | |
| Mean € (95% CI) | 378·43 (308·56 - 448·3) | 297·25 (256·39 - 338·12) | 0·24 |
| Malignant skin lesions | | | |
| Number of persons | 333 | 557 | |
| Mean € (95% CI) | 719·13 (626·88 - 811·38) | 714·69 (647·96 - 781·41) | 0·96 |

Costs are mean costs per person for each subcategory based on the number and types of claims multiplied with costs derived from the open DIS data. P-values were calculated using unpaired two-sided t-tests. CI = confidence interval.

**Table S6.** Characteristics of app-users and non-app-users at 12 months follow-up.

| | **Did not use app** (N = 12,779) | **Used app** (N = 6,230) | **Adjusted p-value** |
|---|---|---|---|
| Age, years | 56·0±14·1 | 53·3±14·1 | <0·001 |
| Sex | | | <0·001 |
| Male | 5607 (43·88%) | 2519 (40·43%) | |
| Female | 7172 (56·12%) | 3711 (59·57%) | |
| Self-reported skin type | | | <0·001 |
| Very white | 5598 (43·81%) | 2825 (45·35%) | |
| White with a beige tone | 6307 (49·35%) | 3075 (49·36%) | |
| Light brown | 784 (6·14%) | 299 (4·8%) | |
| Dark brown | 90 (0·7%) | 31 (0·5%) | |
| Ethnicity | | | 0·002 |
| Dutch | 11694 (91·51%) | 5794 (93%) | |
| European other | 444 (3·47%) | 197 (3·16%) | |
| Non-European | 611 (4·78%) | 226 (3·63%) | |
| Missing | 30 (0·23%) | 13 (0·21%) | |
| Education | | | <0·001 |
| Low | 2326 (18·2%) | 980 (15·73%) | |
| Middle | 4338 (33·95%) | 2304 (36·98%) | |
| High | 5794 (45·34%) | 2818 (45·23%) | |
| Missing | 321 (2·51%) | 128 (2·05%) | |
| Keratinocytic skin cancer in medical history* | 560 (4·38%) | 272 (4·37%) | 0·99 |
| Melanoma in medical history* | 96 (0·75%) | 42 (0·67%) | 0·71 |
| Median number of medications† | 3 (1-5) | 2 (1-5) | 0·001 |

Data are mean (±SD), median (IQR) or n (%). Differences between groups were compared with t-tests for means and medians and $\chi^2$ for categorical variables. IQR = interquartile range. Percentages may not sum to 100 because of rounding.
*Excluding metastases.
† Median number of medications in the year before enrolment in the study, according to the ATC5th level.

**Table S7.** Characteristics of app-users and non-app-users at 24 months follow-up.

| | **Did not use app** (N = 15,825 ) | **Used app** (N = 3,184 ) | **Adjusted p-value** |
|---|---|---|---|
| Age, years | 55·6±14·2 | 52·4±13·6 | <0·001 |
| Sex | | | <0·001 |
| Male | 6,915 (43·7%) | 1,211 (38·03%) | |
| Female | 8,910 (56·3%) | 1,973 (61·97%) | |
| Self-reported skin type | | | <0·001 |
| Very white | 6,965 (44·01%) | 1,458 (45·79%) | |
| White with a beige tone | 7,799 (49·28%) | 1,583 (49·72%) | |
| Light brown | 951 (6·01%) | 132 (4·15%) | |
| Dark brown | 110 (0·7%) | 11 (0·35%) | |
| Ethnicity | | | <0·001 |
| Dutch | 14,517 (91·73%) | 2,971 (93·31%) | |
| European other | 531 (3·36%) | 110 (3·45%) | |
| Non-European | 735 (4·64%) | 102 (3·2%) | |
| Missing | 42 (0·27%) | 1 (0·03%) | |
| Education | | | <0·001 |
| Low | 2,869 (18·13%) | 437 (13·72%) | |
| Middle | 5,571 (35·2%) | 1,071 (33·64%) | |
| High | 6,993 (44·19%) | 1,619 (50·85%) | |
| Missing | 392 (2·48%) | 57 (1·79%) | |
| Keratinocytic skin cancer in medical history* | 675 (4·27%) | 157 (4·93%) | 0·10 |
| Melanoma in medical history * | 98 (0·62%) | 40 (1·26%) | <0·001 |
| Median number of medications † | 3 (1-5) | 2 (1-5) | <0·001 |

Data are mean (±SD), median (IQR) or n (%). Differences between groups were compared with t-tests for means and medians and $\chi^2$ for categorical variables. IQR = interquartile range. Percentages may not sum to 100 because of rounding.
*Excluding metastases.
† Median number of medications in the year before enrolment in the study, according to the ATC5th level.

**Table S8.** Number of histologically confirmed melanomas, cutaneous squamous cell carcinomas and basal cell carcinomas for the participants who did not use the app (n = 12,779) and those who did use the app (n = 6,230) at 12 months follow-up.

| | **Did not use app** (n = 12,779) | **Used app** (n = 6,230) | **Risk Difference (95% CI)** | **Adjusted p-value** |
|---|---|---|---|---|
| Melanoma | 32 | 21 | | |
| Patients with a melanoma | 32 (0·25%) | 21 (0·34%) | 0.09% (-0.09 - 0.27) | 0·36 |
| cSCC | 50 | 33 | | |
| Patients with a cSCC | 46 (0·36%) | 31 (0·5%) | 0.14% (-0.08 - 0.35) | 0·27 |
| BCC | 321 | 170 | | |
| Patients with a BCC | 226 (1·77%) | 137 (2·2%) | 0.43% (-0.01 - 0.87) | 0·10 |
| Total* | 403 | 224 | | |
| Patients with any type of skin cancer* | 292 (2·28%) | 184 (2·95%) | 0.67% (0.16 - 1.17) | 0·03 |

Data are n or n (%) unless otherwise indicated. Differences between groups were compared $\chi^2$ for categorical variables. CI = Confidence interval, cSCC = cutaneous squamous cell carcinoma, BCC = Basal cell carcinoma.
* Only including melanoma, cSCC and BCC.

**Table S9.** Number of histologically confirmed melanomas, cutaneous squamous cell carcinomas and basal cell carcinomas for the participants who did not use the app (n = 15,825) and those who did use the app (n = 3,184) between 12 and 24 months of follow-up.

| | **Did not use app** (n = 15,825) | **Used app** (n = 3,184) | **Risk Difference (95% CI)** | **Adjusted p-value** |
|---|---|---|---|---|
| Melanoma | 37 | 9 | | |
| Patients with a melanoma | 37 (0·23%) | 8 (0·25%) | 0.02% (-0.19 - 0.22) | 1 |
| cSCC | 65 | 8 | | |
| Patients with a cSCC | 58 (0·37%) | 8 (0·25%) | -0.12% (-0.33 - 0.1) | 0·53 |
| BCC | 308 | 98 | | |
| Patients with a BCC | 236 (1·49%) | 75 (2·36%) | 0.87% (0.29 - 1.44) | 0·002 |
| Total* | 410 | 115 | | |
| Patients with any type of skin cancer* | 320 (2·02%) | 90 (2·83%) | 0.81% (0.17 - 1.44) | 0·01 |

Data are n or n (%) unless otherwise indicated. Differences between groups were compared $\chi^2$ for categorical variables. cSCC = cutaneous squamous cell carcinoma, BCC = Basal cell carcinoma.
* Only including melanoma, cSCC and BCC.

**Table S10.** Incidence of participants with a claim for benign and/or (pre)malignant skin lesions split on participant who used the app and who did not use the app at 12 months follow-up.

| | **Did not use app** (N = 12,779) | **Used app** (N = 6,230) | **Risk Difference (95% CI)** | **Adjusted p-value** |
|---|---|---|---|---|
| **Benign skin tumours and nevi** | | | | |
| Dermatology-based claims | 337 (2·6%) | 304 (4·9%) | 2·3 (1·6 - 2·9) | < 0·001 |
| Claims from all relevant specialties | 401 (3·1%) | 340 (5·5%) | 2·4 (1·7 - 3) | < 0·001 |
| **(Pre)malignant skin lesions** | | | | |
| Dermatology- based claims | 819 (6·4%) | 450 (7·2%) | 0·8 (0 - 1·6) | 0·05 |
| Claims from all relevant specialties | 846 (6·6%) | 458 (7·4%) | 0·8 (-0·1 - 1·5) | 0·07 |

Data are n or n (%) unless otherwise indicated. Differences between groups were compared $\chi^2$ for categorical variables. CI = Confidence interval.

**Table 11.** Incidence of participants with a claim for benign and/or (pre)malignant skin lesions split on participant who used the app and who did not use the app between 12 and 24 months of follow-up.

| | **Did not use app** (N = 15,825) | **Used app** (N = 3,184) | **Risk Difference (95% CI)** | **Adjusted p-value** |
|---|---|---|---|---|
| **Benign skin tumours and nevi** | | | | |
| Dermatology-based claims | 369 (2·3%) | 154 (4·8%) | 2·5 (1·7 - 3·3) | < 0·001 |
| Claims from all relevant specialties | 460 (2·9%) | 167 (5·2%) | 2·3 (1·5 - 3·2) | < 0·001 |
| **(Pre)malignant skin lesions** | | | | |
| Dermatology- based claims | 1044 (6·6%) | 238 (7·5%) | 0·9 (-0·1 - 1·9) | 0·08 |
| Claims from all relevant specialties | 1069 (6·8%) | 244 (7·7%) | 0.9 (-0·1 - 1·9) | 0·08 |

Data are n or n (%) unless otherwise indicated. Differences between groups were compared $\chi^2$ for categorical variables. CI = Confidence interval.
